# SARS-CoV-2 Variant Surveillance Using Tandem Targeted RT-PCR-based Genotyping Assays and Whole Genome Sequencing

**DOI:** 10.1101/2022.01.31.22270226

**Authors:** Nicholas P. Pinkhover, Eduardo Sanchez, Kerriann M. Pontbriand, Kenneth Okello, Liam M. Garvey, Kelli P. Fletcher, Alex Pum, Kurvin Li, Gabriel DeOliveira, Teddie Proctor, Jelena D. M. Feenstra, Océane Sorel, Manoj Gandhi, Jared R. Auclair

**Affiliations:** Department of Chemistry and Chemical Biology, Life Sciences Testing Center, Northeastern University Innovation Campus in Burlington, Burlington, Massachusetts; Thermo Fisher Scientific, Genetic Testing Solutions, 180 Oyster Point Blvd, South San Francisco, CA

## Abstract

Genomic surveillance is critical for tracking SARS-CoV-2 Variants of Concern (VOC) and for rapid detection of emerging variants. Whole genome sequencing (WGS) is the predominant method for genomic surveillance; but it is a laborious process for large-scale testing. The aim of this study was to assess the performance of a PCR-based mutation panel for the discrimination of 5 known VOC; Alpha (B.1.1.7), Beta (B.1.351), Gamma (P.1), Delta (B.1.617.2) and Omicron (B.1.1.529). Genotyping analysis was performed on 128 SARS-CoV-2 positive samples collected at the Life Science Testing Center at Northeastern University from April-December 2021. RNA extraction was performed using MagMax™ Viral/Pathogen II Nucleic Acid Isolation Kit. SARS-CoV-2 detection was confirmed using the TaqPath™ COVID-19 Combo Kit. Variant determination was conducted using a panel of TaqMan™ SARS-CoV-2 single nucleotide polymorphism (SNP) assays. On November 25, 2021, the emerging VOC (Omicron) was reported by South Africa and the panel was quickly modified to detect Omicron by substituting P681H and K417N assays. Based on the SNP panel analysis, variant identification in 128 samples were as follows: Alpha (N=34), Beta (N=1), Gamma (N=7), Delta (N=41) and Omicron (N=21). The genotyping panel accurately assigned lineages to 104 samples, confirmed by Ion Torrent GeneStudio S5 WGS. VOC discrimination using RT-PCR genotyping is a rapid, versatile method for detecting known and emerging SARS-CoV-2 variants. The versatility of SNP panels allows monitoring of emerging strains by simple layout adaptations. RT-PCR genotyping assays can expedite variant identification, enable high-throughput variant surveillance, and support WGS prioritization for detection of new variants.

## Introduction

Severe acute respiratory syndrome coronavirus 2 (SARS-CoV-2), the causative agent of coronavirus disease 19 (COVID-19) was discovered in Wuhan, China in December of 2019. From poor disease monitoring programs to inconsistent public health precautions, numerous factors impacted the evolution of the COVID-19 pandemic.^1,2^ Vaccination coverage, non-pharmaceutical interventions and surveillance testing strategies are critical for mitigating the impact of COVID-19 but as these protective measures vary across countries SARS-CoV-2 continues to spread worldwide.^1-3^

Since September 2020 several variants of SARS-CoV-2 have emerged that have been labeled variants of concern (VOC) or variants under monitoring (VUM) based on their properties such as increased transmissibility, virulence, or impact on the performance of therapeutics, vaccines, or diagnostic tests.^1-7^ Alpha (B.1.1.7), Beta (B.1.351) and Gamma (P.1) VOC circulated in the first half of 2021, before the emergence of Delta (B.1.617.2) VOC replaced Alpha as the dominant variant in July 2021.^3,5,6^ Recently a new VOC, Omicron (B.1.1.529), has emerged and currently accounts for 99.5% of all SARS-CoV-2 cases in the United States.^3^ Sequence analysis of VOC revealed the S-gene as a site bearing numerous mutations, some of which are shared among different VOC.^3-6,8^ This finding highlights the important need of a genotyping approach for tracking mutations of concern (MOC) expressed in SARS-CoV-2 variants. Indeed, disease monitoring programs should exhibit high sensitivity and quick turn-around-times, while workflows should remain largely unaffected by varying positivity rates. Here, we describe an economic, convenient, and multimodal method for SARS-CoV-2 genomic surveillance using targeted RT-PCR-based genotyping assays (Table 1).

**Table 1.**
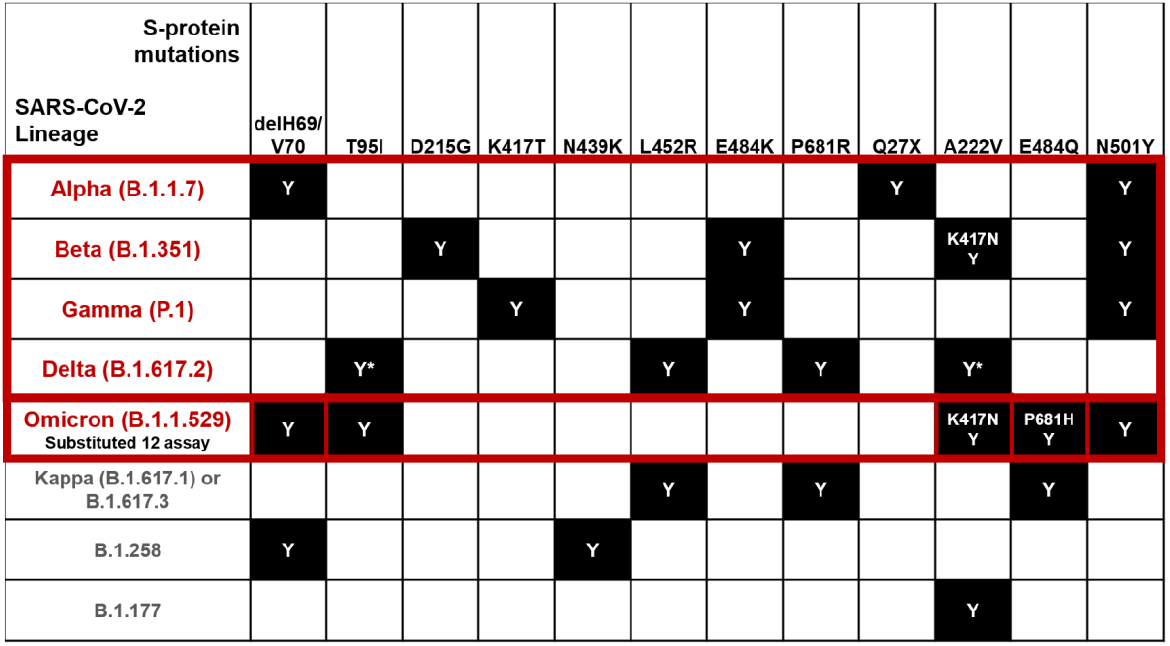
Panel of mutations in the SARS-CoV-2 S-protein selected for genotyping and combinations of mutations used for lineage assignment to five variants of concern: Alpha, Beta, Gamma, Delta, and Omicron (in red) and other variants of interest/under monitoring (in grey).

The Life Science Testing Center (LSTC) at Northeastern University (NU) is a CAP and CLIA certified clinical laboratory established in August 2020 for routine COVID-19 testing for NU’ s student, faculty, and staff. As of January 22, 2022, 1,411,171 samples have been analyzed using RT-PCR of which, 7328 samples have tested positive, mirroring the Greater Boston positivity rates and trends^9^ (Figure 1A). In this study, an RT-PCR-based genotyping approach was developed and implemented to conduct extensive VOC screening. We analyzed N=128 confirmed SARS-CoV-2 positive samples collected between April and December 2021 thus covering the period when Alpha, Beta, Gamma, Delta, and Omicron VOC were circulating.^3,8,10^ In addition, vaccine breakthrough cases (N=226) were tested between July and December 2021 using this targeted RT-PCR-based genotyping approach.^11-15^ In this study the analytical performance and economic impact of a targeted genotyping approach was compared to whole genome sequencing (WGS) for large-scale screening of SARS-CoV-2 VOC.^5,6,8^

**Figure 1.**
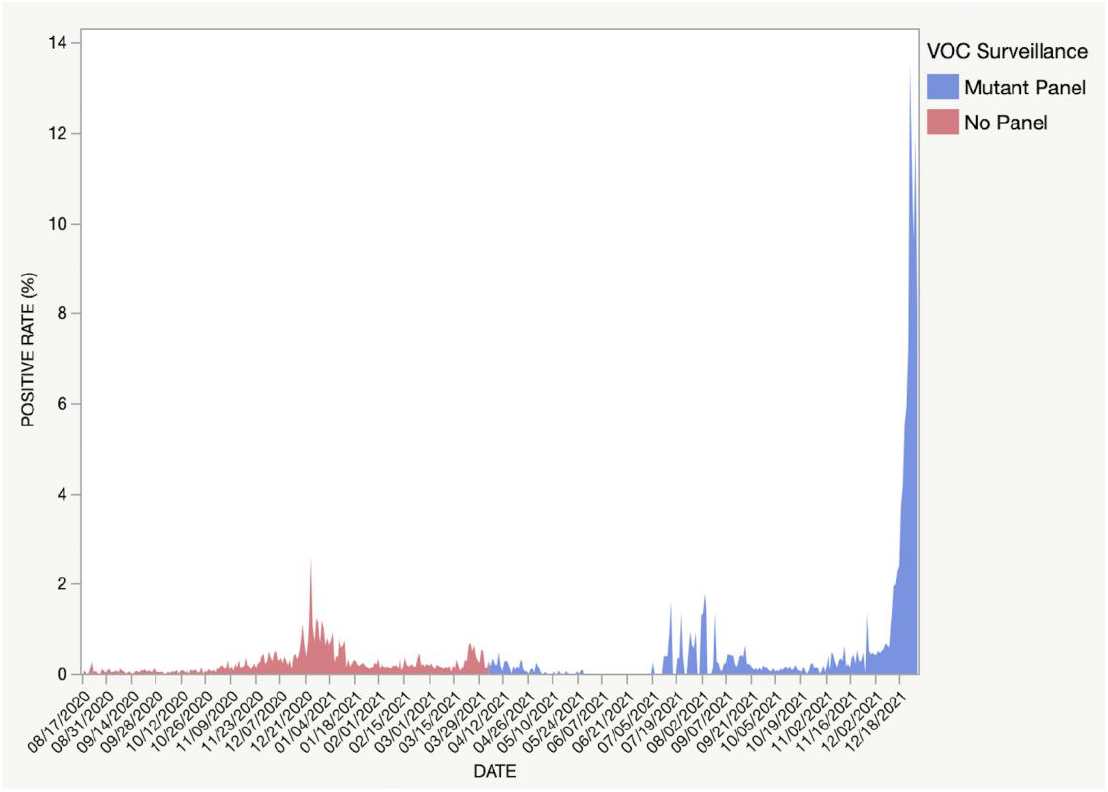
**A**. SARS-CoV-2 test positivity rate in LSTC population between August 2020 and December 2021. Samples that were tested using the mutation panel are highlighted in blue whereas samples that were tested before implementation of our VOC surveillance assay are depicted in red.

## Results

A total of 128 SARS-CoV-2 positive samples that were initially collected between April and December 2021 were tested using the targeted RT-PCR-based genotyping assay. This RT-PCR approach included either an 8-assay (N=44 samples tested) or a 12-assay (N=84 samples tested) MOC panel (Table 1) to detect SARS-CoV-2 VOC and VUM. Using the interpretation criteria described in Table 1, samples were assigned to the following SARS-CoV-2 lineages: Alpha (B.1.1.7; N=34), Beta (B.1.351; N=1), Gamma (P.1; N=7), Delta (B.1.617.2; N=41) and Omicron (B.1.1.529; N=21). The 8-assay panel defined SARS-CoV-2 lineages to 21 of 44 (47.72%) total samples tested, and the 12-assay panel defined SARS-CoV-2 lineages to 83 of 84 (98.88%) total samples tested within the screened positive sample pool (N=128), allowing for a combined mean performance of 81.25% discrimination of SARS-CoV-2 VOC lineages (Figure 1B). Overall, 24 SARS-CoV-2 positive samples were not assigned to any VOC lineage, however, it should be noted that 39 samples of the 128 tested were pooled-swab surveillance samples. Eight of the 24 samples with undetermined VOC status were pooled samples, which infringes on the intended application of the mutation panel (see SI). The Alpha VOC was most pronounced in April-May 2021, while the Delta variant was detected over the entire screening period with a peak in cases in August (Figure 1B). In April 2021, we detected a spike in non-VOC positive samples which could be attributed to unstable viral recombination prior to Delta’ s stabilization (Figure 1B).^10,12,13^ Finally, the Omicron variant became the most frequently detected VOC in December 2021 (Figure 1B).^3^ No significant difference in mean Ct values for any of the three gene targets reported by the TaqPath™ COVID-19 Combo Kit used for SARS-CoV-2 detection were observed between the 5 VOC (see SI).^14^

**Figure 1.**
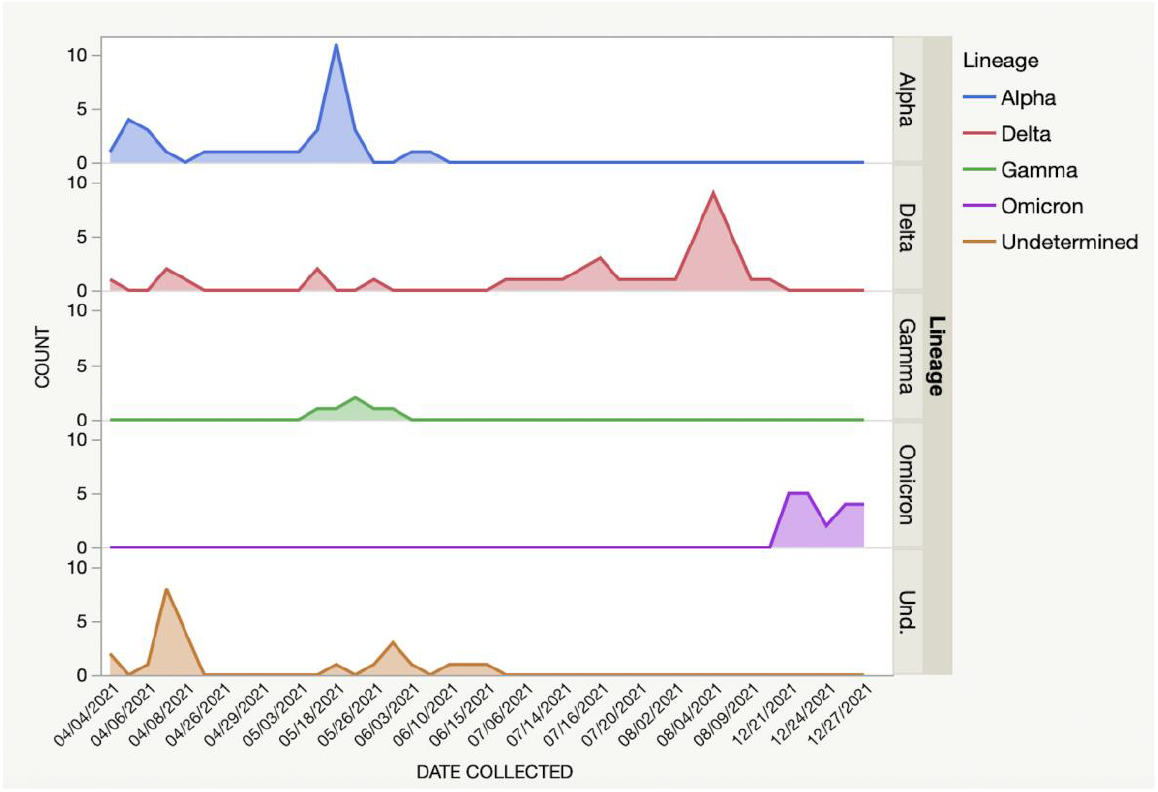
**B**. Frequency of identified/undetermined SARS-CoV-2 lineages in SARS-CoV-2 positive specimen collected from April to December 2021 using targeted RT-PCR based genotyping assay.

In addition, VOC trends were monitored in vaccine breakthrough COVID-19 cases from July-December 2021, where breakthrough cases (N=226) in fully vaccinated (two-dose mRNA) individuals were recorded, respectively (Figure 1D). The higher frequency of breakthrough cases coincided first with the broad wave of Delta cases (N=151) from July to December 2021^10,12-15^, followed by a rapid spike of Omicron cases (N=75) December 01 to December 27, 2021, at Northeastern University, as identified by the MOC panel genotyping approach (Figure 1D).

A side-by-side cost analysis was performed by comparing the mutation panel genotyping approach for VOC surveillance to WGS, with respect to cost of instrumentation, consumables, staff, computing capacity and data storage and other variables (Figure 1C).^4-6,8^ The average cost per sample for the genotyping approach using a 12-assay mutation panel was calculated to be over 8 times lower than WGS, without factoring in the longer turn-around-time for obtaining the WGS results when compared to RT-PCR.^5,6,8^

**Figure 1.**
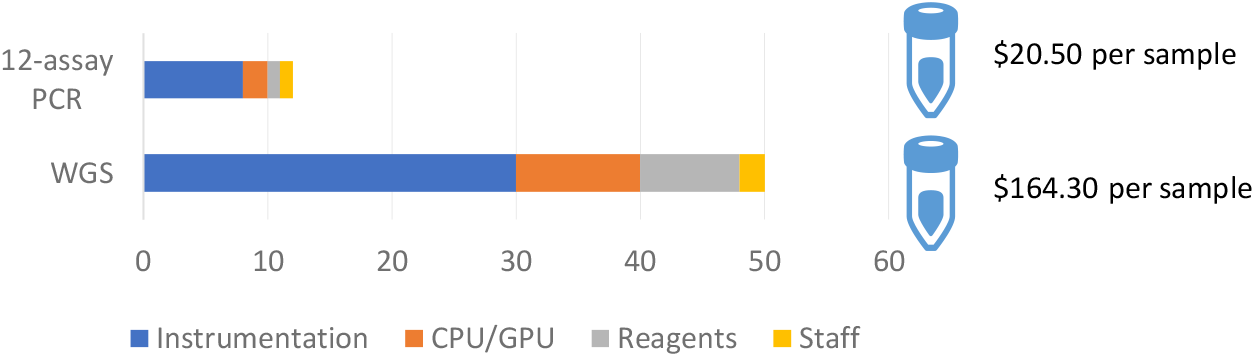
**C**. Cost benefit analysis between 12-assay Mutant Panel and WGS for variant of concern surveillance.

**Figure 1.**
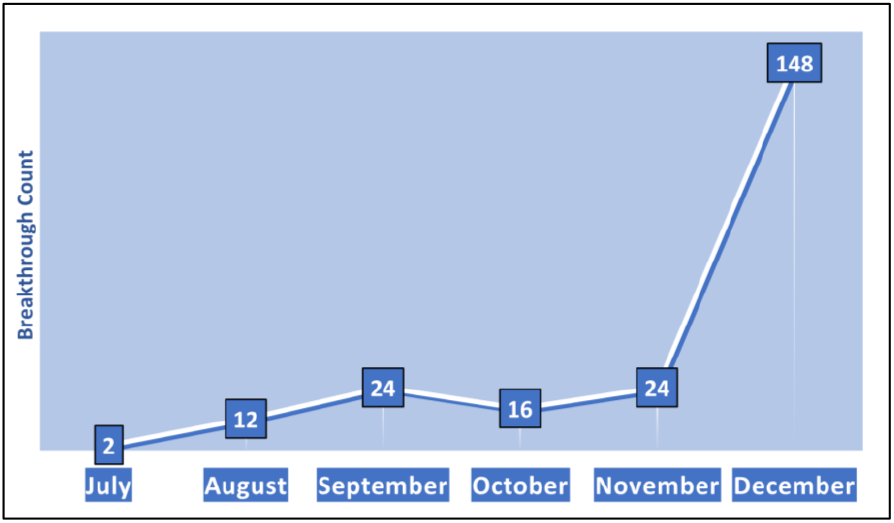
**D**. Breakthrough SARS-CoV-2 positive incidence (N=226) in fully vaccinated (14-days after second dose of either Moderna or Pfizer COVID-19 mRNA vaccine) patient samples tested by the LSTC after Northeastern began its COVID-19 vaccination program on January 05, 2021. A distinct spike in breakthrough infection in December 2021 was notably correlated with the emergence of the Omicron VOC on NU’ s campuses, as Delta (N=73) and Omicron (N=75) accounted for all 148 of December breakthrough cases.

## Discussion

Surveillance of SARS-CoV-2 variants is essential for timely implementation of public health measures aimed at limiting viral spread and effects of the ongoing COVID-19 pandemic.^3,11-15^ While WGS has proven to be an invaluable tool for identification of new variants, the cost as well as turnaround times for sample processing and data analysis remain challenging for wide implementation.^4-6,8^ This demonstrates that genotyping assays offer a fast, reliable, and cost-effective method for monitoring of known SARS-CoV-2 variants within a population.

Results showed that mutation panels were able to accurately assign lineages to VOC as confirmed with WGS. Although genotyping assays can only assign samples to lineages that are already defined in the database, a key feature of genotyping assays is that they can be rapidly updated as new VOC exhibiting new combinations of mutations arise^4-6^, which can be powerful given the rapid emergence and spread of previous VOC.^3,10-12,14^

This study showed that when comparing surveillance technical costs, the 12-Assay Mutation panels afforded 98.88% VOC determination at >8 times cheaper than WGS testing per sample, and costs can be further reduced by targeting selected VOC. Importantly, this targeted RT-PCR-based genotyping approach enables high-throughput variant surveillance and can be easily implemented in a routine testing laboratory including in developing regions with limited access to WGS. With minimal automation, this genotyping approach generated VOC data for 96 samples with a turnaround of ∼3.5 hours whereas WGS required 5-7 days to confirm VOC data on 96 samples. Thus, expanding the use of a RT-PCR-based mutation panel approach would complement WGS for tracking emergence and surveillance of VOC. WGS could be prioritized to target samples that have not been assigned to a VOC lineage using mutations panels or that are consistently inconclusive in a routine SARS-CoV-2 RT-PCR test.

The use of mutation panels can further be valuable to measure the impact of VOC on parameters such as vaccine effectiveness in preventing SARS-CoV-2 infections. Data showed a strong association between the emergence of both Delta and Omicron variants and an increase in the number of breakthrough infections during the same periods. Rapid tracking of variants with immune escape or higher transmissibility is key as COVID-19 transitions into an endemic state.

Taken together, these results show that VOC genotyping using mutations panels is a compelling and versatile approach to monitor variant prevalence in real-time and that should be leveraged to adjust public health strategy.

## Materials and Methods

### Identification of Positive SARS-CoV-2 samples

This study included anterior nasal swabs specimens collected between April and December 2021 at NU. Viral RNA extraction and purification was performed using the MagMax™ Viral/Pathogen II Nucleic Acid Isolation Kit and RT-PCR with TaqPath™ COVID-19 Combo Kit (both Thermo Fisher Scientific) on Applied Biosystems™ 7500 Fast Dx RT-PCR thermocyclers. 128 SARS-CoV-2 positive samples representing pooled (N=39) samples, asymptomatic (N=43) and symptomatic (N=26) individuals were subjected to variant of concern identification through target RT-PCR-based genotyping.

### Targeted RT-PCR-based Genotyping

SARS-CoV-2 VOC were detected using TaqMan™ SARS-CoV-2 Mutation Panel (Thermo Fisher Scientific) in an 8-assay or 12-assay format as defined in Table 1. Data analysis was performed using Applied Biosystems™ Design and Analysis Software Version 2.5.1.

For details refer to SI.

## Data Availability

All data produced in the present study are available upon reasonable request to the authors

https://assets.thermofisher.com/TFS-Assets/LSG/manuals/MAN0024768_TaqManSARS-CoV-2_MutationPanel_UG.pdf

https://assets.thermofisher.com/TFS-Assets/LSG/manuals/MAN0019746_MagMAX_MVP_II_NA_IsolationKit_CE-IVD_IFU.pdf

https://assets.thermofisher.com/TFS-Assets/LSG/manuals/MAN0019181-RevK-TaqPathCOVID19Kit-IFU-EUA.pdf

https://assets.thermofisher.com/TFS-Assets/LSG/manuals/MAN0019277_Ion_AmpliSeq_SARS-CoV-2_Research_Panel_GeneStudio_QR.pdf

## Acknowledgments

Thank you to all involved in making this research possible. We are grateful for the incredible dedication, time, and energy provided by everyone at the LSTC to complete this impactful project.

