## Supplementary Information for "SARS-CoV-2 Variant Surveillance Using Tandem Targeted RT-PCR-based Genotyping Assays and Whole Genome Sequencing"

### Extended Methods

### Identification of Positive SARS-CoV-2 samples

Clinical determination of positive SARS-CoV-2 status was carried out by testing anterior nasal swabs collected at the Cabot Testing Center and processed by the Life Sciences Testing Center at Northeastern University. Viral RNA extraction and purification were performed using the Applied Biosystems™ MagMax™ Viral/Pathogen II Nucleic Acid Isolation Kit (Thermo Fisher Scientific) on the Agilent™ Bravo automated Liquid platform (Agilent Technologies, Inc., Santa Clara, CA 95051). Following RNA isolation, RT-qPCR was conducted using Applied Biosystems™ TaqPath™ COVID-19 Combo Kit (Thermo Fisher Scientific) and the Applied Biosystems™ 7500 Fast Dx Real -time PCR system (Thermo Fisher Scientific).

### Variant of Concern Identification

TaqMan™ SARS-CoV-2 Mutation Panel (Thermo Fisher Scientific) was used for the detection of variants of concern (VOC) and additional SARS-CoV-2 variants (Table 1). Viral RNA was extracted and purified from original clinical samples using the Applied Biosystems™ MagMax™ Viral/Pathogen II Nucleic Acid Isolation Kit on Agilent™ Bravo automated Liquid platform. Extraction/purification was conducted following a modified TaqPath™ FDA-EUA extraction protocol, where MS2 phage control was not added to samples to prevent interference during single nucleotide polymorphism (SNP) genotyping using the mutation panels.

RNA samples were processed according to the standard TaqMan™ SARS-CoV-2 Mutation Panel Assay protocol. RT-PCR was then conducted on a QuantStudio™ 6 RT-PCR system (Thermo Fisher Scientific) following the recommended TaqMan™ SARS-CoV-2 Mutation Panel thermal and PCR settings.. Data analysis was performed using Applied Biosystems™ Design and Analysis Software Version 2.5.1 (Thermo Fisher Scientific).

Variant genotyping determination was carried out using allelic discrimination displayed as scatter plots. SNPs from samples expressing copies of (FAM-labeled) mutant alleles displayed linear amplification in the y-axis, while reference sequence (VIC-labeled) samples clustered linearly along the x-axis. Further statistical and data visualization on all collected data was performed using JMP® Pro 15.0.0, and Microsoft® Excel® for Microsoft 365 MSO. Mutation panel results and assignment to VOC were verified by Whole Genome Sequencing using Thermo Fisher Scientific's Ion Torrent- GeneStudio-S5 Plus System.

### Data Set S1: ANOVA of TaqPath™ Cycle Threshold (Ct) Values in VOC

| Lineage | Sample Size (N= 125) | Ct Mean [95% CI] | | |
|---|---|---|---|---|
| | | ORF1ab-Gene | N-Gene | S-Gene[a] |
| Alpha | 34 | 22.323 [20.688, 23.958] | 22.4479 [20.864, 24.032] | N/A |

| | | | | |
|---|---|---|---|---|
| Delta | 41 | 23.2902 [21.801, 24.779] | 23.8917 [22.449, 25.334] | 23.6217 [21.703, 25.54] |
| Gamma | 5 | 27.1976 [22.933, 31.462] | 27.8692 [23.738, 32] | 28.5291 [22.854, 34.204] |
| Omicron | 20 | 17.6305 [15.498, 19.763] | 18.6984 [16.633, 20.764] | N/A |
| Und. | 24 | 22.3755 [20.429, 24.322] | 22.7259 [20.84, 24.612] | 22.2285 [15.675, 28.782] |
| One Way ANOVA | P-Value | 0.0001 | 0.0002 | 0.2289 |

[a]Calculated Ct means excluded positive samples that did not detect the S-gene target. The adjusted N after exclusion for the lineages were Delta (N= 35), Gamma (N= 4), and Und. (N= 3). Alpha and Omicron lineages were excluded entirely due to total S-gene target failure (SGTF).

**SI References**

Sample References:


1. https://assets.thermofisher.com/TFS-Assets/LSG/manuals/MAN0024768_TaqManSARS-CoV-2_MutationPanel_UG.pdf
